# Rapid Detection of SARS-CoV-2 Antibodies Using Electrochemical Impedance-Based Detector

**DOI:** 10.1101/2020.08.10.20171652

**Authors:** Mohamed Z. Rashed, Jonathan A. Kopechek, Mariah C. Priddy, Krystal T. Hamorsky, Kenneth E. Palmer, Nikhil Mittal, Joseph Valdez, Joseph Flynn, Stuart J. Williams

## Abstract

Emerging novel human contagious viruses and pathogens put humans at risk of hospitalization and possibly death due to the unavailability of vaccines and drugs which may take years to develop. Coronavirus disease (COVID-19) caused by severe acute respiratory syndrome coronavirus 2 (SARS-CoV-2) was classified as a pandemic by the World Health Organization and has caused over 550,000 deaths worldwide as of July 2020. Accurate and scalable point-of-care devices would increase screening, diagnosis, and monitoringof COVID-19 patients. Here, we demonstrate rapid label-free electrochemical detection of SARS-CoV-2 antibodies using a commercially available impedance sensing platform. A 16-well plate containing sensing electrodes was pre-coated with receptor binding domain (RBD) of SARS-CoV-2 spike protein, and subsequently tested with samples of anti-SARS-CoV-2 monoclonal antibody CR3022 (0.1 μg/ml, 1.0 μg/ml, 10 μg/ml). Subsequent blinded testing was performed on six serum specimens taken from COVID-19 and non-COVID-19 patients (1:100 dilution factor). The platform was able to differentiate spikes in impedance measurements from a negative control (1~ milk solution) for all CR3022 samples. Further, successful differentiation and detection of all positive clinical samples from negative control was achieved. Measured impedance values were consistent when compared to standard ELISA test results showing a strong correlation between them (R2 = 0:9). Detection occurs in less than five minutes and the well-based platform provides a simplified and familiar testing interface that can be readily adaptable for use in clinical settings.

## 1. Introduction

The recent emergence of novel coronavirus disease (COVID-19) is caused by the severe acute respiratory syndrome coronavirus 2 (SARS-CoV-2). It was classified as a world pandemic by the World Health Organization (WHO) due to its rapid human to human transmission nature with an estimated basic reproduction number of 2.2 (Li et al. (2020)). There are ongoing efforts to develop drug treatments and vaccines for COVID-19, but until reliable management of this disease is implemented there is a considerable need to develop rapid diagnostic tests that are readily available and can be integrated with current healthcare protocols to in order to monitor the spread of the virus.

Immunoassays detect binding of antibodies to a specific target molecule and can be used for detecting the presence and concentration of specific antibodies within an infected patient. Antibody screening has several advantages including to determine if a patient has been previously infected, evaluate the prevalence of the disease, identify candidates for convalescent plasma therapy, and assess the immune response induced by a vaccine. The standard diagnostic method for precise and quantitative antibody detection for most common pathogen including SARS-CoV-2, is the enzyme-linked immunosorbent assay (ELISA) (Law et al. (2015)); however, this detection method can take hours to perform due to sample processing and incubation steps.

Over the past few decades, biosensors have emerged as a powerful tool to complement ELISA methods. Biosensors are the integration of a biorecognition element which remains in contact with the target molecule and a transduction element that confirms target binding and detects the event with a measurable electrical signal. Biosensors may provide qualitative or quantitative information, many without the need for an additional pre-labeling or tagging process. A variety of biosensing platforms have been developed for respiratory viral infections (Saylan et al. (2019)) and COVID-19 (Choi (2020)). There are many types of biosensors for antibodies (Liu and Jiang (2017)), many rely on optical, chemical, and/or electrical mechanisms to produce a measure-able signal. The method discussed herein is electrochemical impedance-based sensing (EIS) using commercially available equipment, though its sensitivity can likely be improved with modifications.

EIS detection methods are cost-effective, typically label-free, and enable high precision for detection of antibodies or pathogens (Cesewski and Johnson (2020); Leva-Bueno et al. (2020)). EIS measures the changes in applied electric current or voltage due to the binding of the target molecule. Al-ternatively, frequency analysis response methods, referred to as impedance-based analysis methods, measure the changes in the phase angle and impedance between the solution and working electrode as a function of the frequency of the applied voltage/current (~ 3 kHz - 10 MHz). Here, an equivalent circuit (containing combination of resistors, capacitors, and capacitive phase elements) is modeled after the sensing region and the measured impedance and phase angle data is used to extract values of the fit circuit elements to monitor changes in system behavior. Capacitive elements typically represent the behavior of the ionic double layer at the electrode-electrolyte interface, which can provide essential information about the nature and rate of probe-target binding. Even though measuring the impedance across wide range of frequencies is useful for comprehensive biosensing applications, fixed or single frequency measurements can provide improved transient information about rapid changes in double-layer capacitance due to increasing sampling. Also, operating at fixed frequencies simplify EIS platform hardware, eliminating the need for complex frequency generators and enabling portable analysis.

Capacitive immunosensing is an emerging EIS technique that focuses on sensing changes in the electric double layer (EDL) through sensing binding events between the probe immobilized on the electrode surface and suspended target (Berggren et al. (2001)). The performance of capacitive immunosensors can be adjusted by modifying the electrode surface area, insulation (coating) layer, and the electrode and coating material. Highly sensitive sensors can be achieved by making thin conformal coating layer which ensure stable capacitance changes in response to probe/target binding (Castiello et al. (2019)). Several studies reported the use of capacitance immunosensors for detection of pathogens (Cesewski and Johnson (2020); Hanna et al. (2006); Luka et al. (2019); Teeparuksapun et al. (2012); Wang et al. (2019, 2017)), thus demonstrating that it can be used as a reliable label-free sensing method.

In this study, we report a capacitive immunosensing assay using a commercially-available impedance detection system that uses specialized well-plates that have integrated sensing electrodes from ACEA Biosciences (Figure 1). ACEA Biosciences’ 16-well plate xCELLigence system (RTCA S16) design was intended for non-invasive EIS detection of cell proliferation, morphology change, and attachment quality. Each well contains an array of specially-designed interdigitated electrodes fused to polyethylene terephthalate. The plate interface independently addresses each well to acquire single frequency measurements (10 kHz) every few seconds. Here, we demonstrate that it can be used to successfully detect SARS-CoV-2 antibodies. We hypothesize that the detected mechanism corresponds to binding kinetics between the SARS-CoV-2 spike protein receptor binding domain (RBD) and the anti-SARS-CoV-2 antibody. This approach provides quick and reliable results, is relatively compact and portable, and has the potential to be scaled up (simultaneous scanning of up to four 384 well plates is possible); thus, this approach is a good candidate for rapid screening of serum samples for SARS-CoV-2 antibodies.

**Figure 1:**
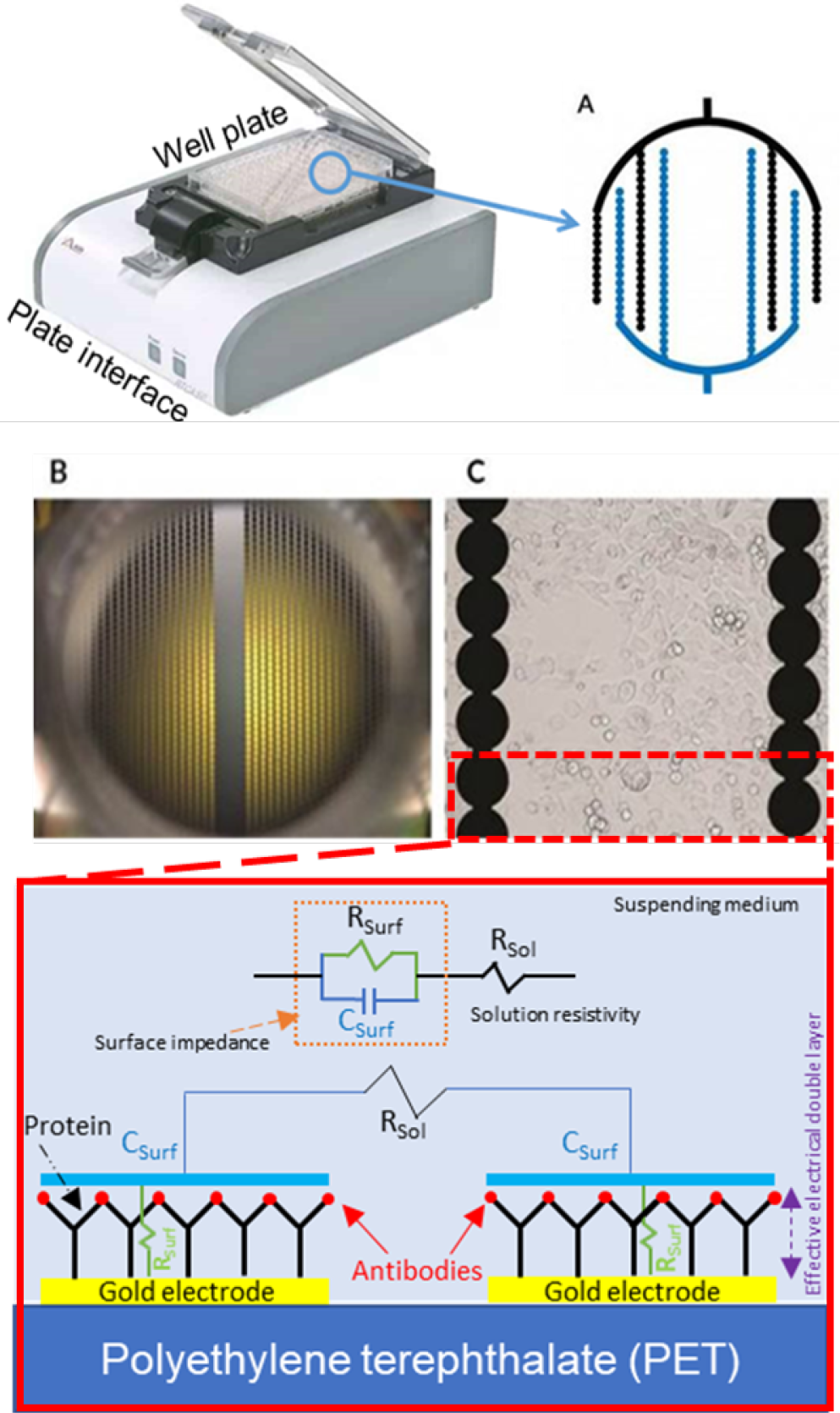
(A) Image of ACEA Bioscience’s 96-well platform with a (B) schematic of the electrode layout. (C) Actual image of the electrodes themselves within the well and a (D) magnified image of the electrodes. Images A-D are modified from ACEA Bioscience. (E) Schematic of electrical impedance equivalent circuit model of the protein/antibody in solution.

## 2. Methods

Figure 1 shows ACEA Bioscience’s well platform with its integrated electrodes. The specialized well-plate is attached to an interface and is connected via USB to a laptop. Software controls the timing of measurements for each independently addressable well. Impedance measurements with respect to time, *Z*(*t*), were acquired continuously at 10 kHz and 22 mV. Results herein are reported as a change in impedance, ∆*Z* = *Z(t) − Z_o_*, with respect to a measured control, *Z_o_*. The impedance model in Figure 1E depicts the mechanism of the spike RBD antigen proteins, medium solution, and anti-SARS-CoV-2 antibodies. The total impedance includes the surface impedance which combines both EDL capacitance, *C_Surf_*, in parallel with double layer resistance, *R_Surf_*. The surface impedance is in series with the ionic medium resistance, *R_Sol_*, which arises from the conductivity of the ions in the solution and is not altered by the binding between probe protein and target antibodies. For simplicity, we are neglecting any insulative effects that may be present on the electrode surface and their effect on the total impedance. The total measured impedance, *Z(t)*, is a function of in the EDL resistance and capacitance described with (Quoc et al. (2017)):

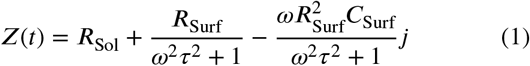

where *τ* = *R*_Surf_*C*_Surf_. From Eq. (1), the series capacitances term, *C_s_*, can be extracted from the imaginary part of the impedance as the following:

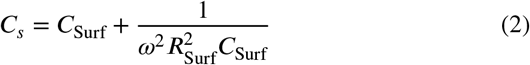

As can be seen from Eq. (2), Cs is frequency dependent. At high frequencies, it will converge to *C*_Surf_, however, at low frequencies, *C_s_* will be dependent on, *C*_Surf_, and *R*_Surf_. Hence, protein/antibody binding events will influence the EDL resistance and capacitance at low frequencies (?) and, thus, 10 kHz is an appropriate sensing frequency for this study.

To initially validate our approach, a monoclonal anti-SARS-CoV-2 antibody (CR3022, Abcam, Cambridge, MA, USA) was tested. The well plates were coated with 2.5 μg/mL SARS-CoV-2 spike RBD protein (obtained from BEI Resources, NR-52306) diluted in PBS, 50 μL/well was added, and plates were sealed and refrigerated at 4 °C for 1-3 days. Next, wells were decanted and washed with 200 μL of 0.1% PBS/Tween-20 (PBS-T) solution three times using manual pipetting. To perform the test, the wells were filled with 100 μL of blocking solution (3% milk) and incubated for 1 hour. The measurement wells were decanted and 100 μL of 1% milk solution buffer (background buffer) was added to each well, then a background measurement (*Z_o_*) was acquired. Next, half of the background buffer was removed (50 μL) and 50 μL of the sample solution containing CR3022 antibody (0.1 μg/ml, 1.0 μg/ml, or 10 μg/ml) diluted in 1% milk solution was applied to the well while simultaneous impedance measurements were continuously acquired.

Tests with six human serological samples were provided (Serum samples were collected following protocols approved by the University of Louisville Institutional Review Board) and the impedance team was blinded to clinical information about the samples *(i.e*., whether they were positive or negative for anti-SARS-CoV-2 antibodies as validated by ELISA). Wells were prepared and background impedance measurements were acquired as previously described for the CR3022 tests. Next, 50 μL was aspirated from each well and replaced with a 50 μL of human serological samples (1:100 dilution factor) while continuous impedance measurements were acquired. Impedance measurements were compared to ELISA measurements on the same serological samples. ELISA was performed by coating immunoplates with of SARS-CoV-2 spike RBD protein overnight, washing three times in PBS-T, blocking for 1 hr, incubating for 30 min with human serological samples diluted 1:100, washing 3 times in PBS-T, and incubating 1 hr with HRP-labeled secondary anti-human IgG antibody. To perform the peroxidase reaction, wells were washed three times, TMB solution was added for 3 min, the reaction was stopped by addition of 1M HCl, and optical density of each well was measured on a platereader.

Impedance measurements were exported and analyzed further in MATLAB. Impedance peaks were identified as measurements that increased at least ten times greater than background noise oscillations (>78 mΩ) when compared to its baseline value. The action of adding 50 μL buffer, as will be shown later, generated an increase in impedance of approximately 50 mΩ, which is less than our specified detection criteria. The reported peak value is the mean of the first recorded peak value and the subsequent four impedance measurements. The time between impedance measurements for the same well was approximately two seconds.

## 3. Results and Discussion

Figure 2A shows a representative impedance curve for each tested CR3022 dilution. Figure 2B shows the reported average peak value for each dilution (n = 4 wells each). The addition of CR3022 antibody to the RBD-coated wells caused a rapid increase in measured impedance followed by a decay during the next few minutes. There is inherent error associated with the measured maximum peak value due to the timing of the recorded impedance measurements; greater sensitivity and precision is possible if data acquisition for each well occurred with a period faster than the tested two-second intervals. To demonstrate the need for faster transient measurements, we measured the addition of CR3022 (10 μg/ml) and a control *(i.e*., decanting and adding 50 μL of buffer) with continuous impedance acquisition using a different impedance analyzer system (Agilent 4294A at 10 kHz). Results (Supplemental Figure s1) demonstrated that the measured peak rapidly decreases in less than one second before subsequently decaying more slowly; thus, increased sensitivity and accuracy is possible at higher temporal resolution.

**Figure 2:**
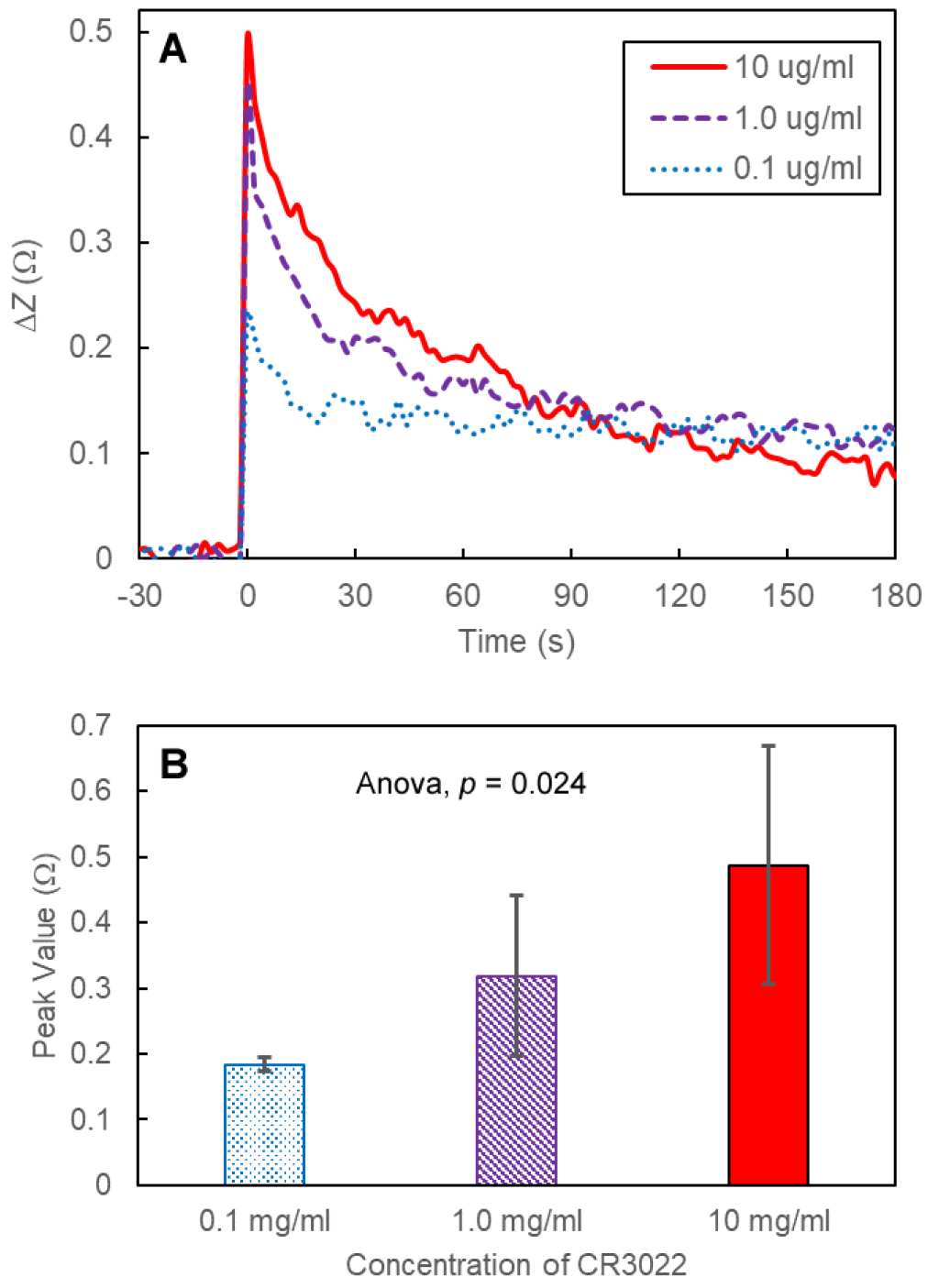
(A) Four arbitrary experimental curves were selected to demonstrate a representative response for each tested concentration and a negative control. (B) Average measured peak impedance values for each concentration (n = 4); error bars represent +/− one standard deviation.

Figure 3A shows a representative impedance curve for two positive and one negative serological sample. The positive impedance signature resembles closely to the CR3022 measurements in that there was a sharp peak followed by a slower decay. The two positive serological samples were from two patients with different antibody levels; similar to the CR3022 levels, impedance spectroscopy can be used to quantify antibody levels in serological samples. Figure 3B compares the measured impedance peak value to ELISA levels. These results show a clear correlation between impedance values and the concentration of antibodies within the sample. The rapid changes in total impedance in our experiments is due to the association and disassociation of the protein/antibody which is happening within few seconds after the introduction of the sample (Kulin et al. (2002)). Dipoles in the bound antigen headgroup can contribute to the measured capacitance because they affect the dielectric constant *(∊_r_)* of the surface impedance. During the binding event, the dielectric constant of the surface layer drops from *∊_r_* ≈ 80*∊*_0_ (for PBS) to *∊_r_* ≈ 2 − 5*∊*_0_ (Daniels and Pourmand (2007); Sondag-Huethorst and Fokkink (1995); Świetlow et al. (1992)) which subsequently decreases the capacitance and, therefore, increases the overall measured system impedance. It’s noted that these dipoles would not react or contribute to the total impedance if the applied signal frequency exceeds 1 MHz (Bordi et al. (2004); Feldman et al. (2003)), which rationalize the operation at a low signal frequency (10 kHz).

**Figure 3:**
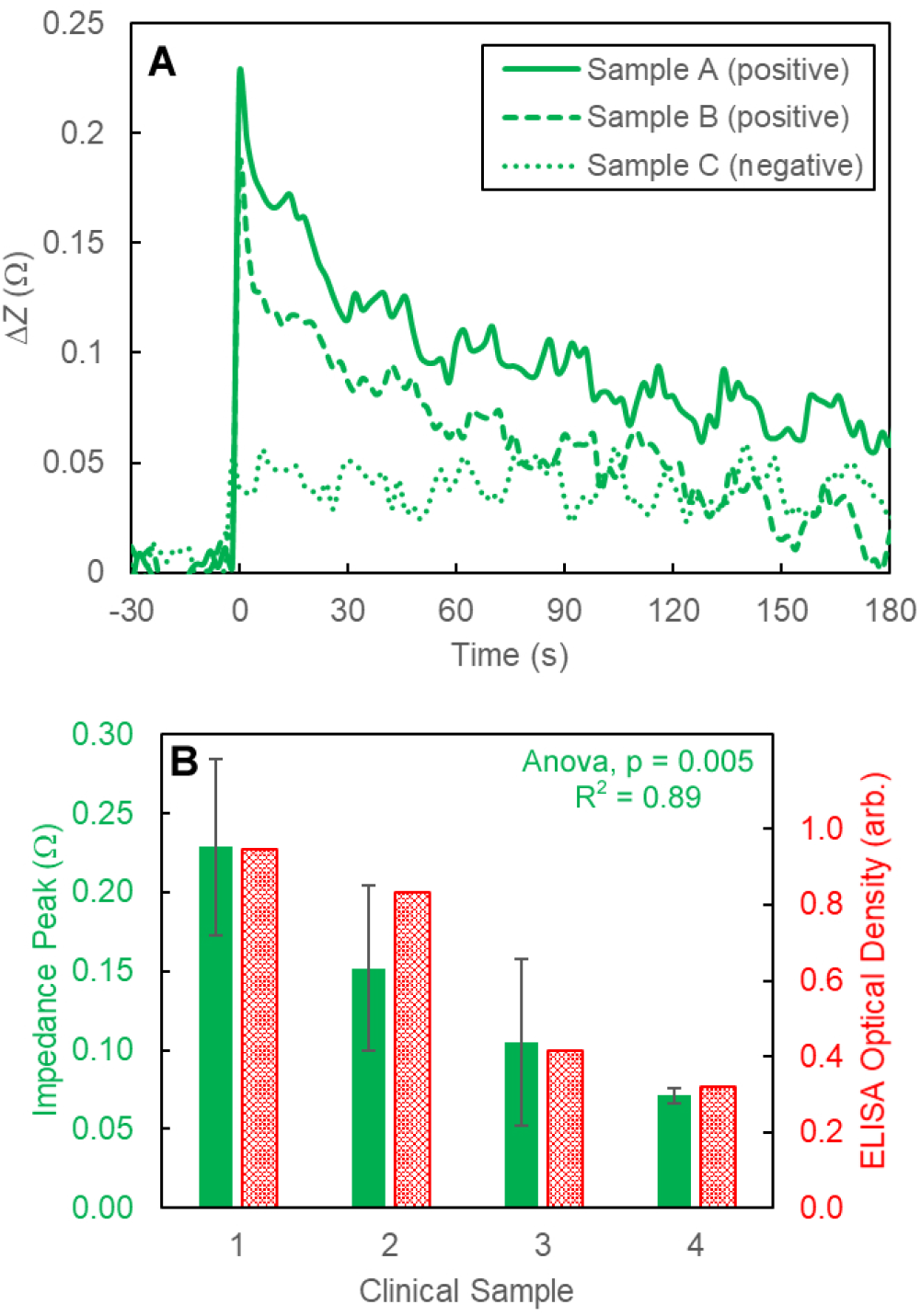
(A) A representative impedance spectra curve for a positive and negative clinical sample. (B) Comparison between the measured impedance peak value (green) when compared to ELISA levels (red).

The limit of detection using this approach is limited by hardware noise and variations in sample handling. Although the lower limit of detection was not demonstrated here, these experiments demonstrate that this implementation of EIS can detect clinically relevant antibody concentrations. The ability to automize the system can offer high throughput operation. Impedance systems also lend themselves to be portable, including potential smartphone-based analytical systems. Further improvements can be made to enhance sensing and throughput of the platform. For example, decreasing the spacing between interdigitated electrodes will reduce the influence of Rsus. Also, electrodes with a thin conformal coating of insulative material would enhance the stability and sensitivity of the capacitance measurement (Castiello et al. (2019)). Future tests will determine if measurements acquired at other frequencies and/or voltages (*i.e*., other than 10 kHz, 22 mV) will further improve platform performance. There are alternative electrochemical detection schemes that could be used for antibody detection. Field-effect transistor-based biosensors (Gaurav and Shukla; Seo et al. (2020)) offer a good approach for detection of the antibodies through the induced variations in source-drain channel conductivity that arise from the electric field of the sample environment due to the binding of the target with the bioreceptors immobilized on the metal/polymer gate. However, such platforms are either expensive, require complicated fabrication methods, or need sophisticated equipment to operate. Further, a limitation in applying FET-based sensors is that their performance may be critically obstructed by the presence of multiple ligands and proteins in serum samples. Moreover, in terms of electrical detection, the detection of biomolecules is severely hindered by ionic screening effect caused by high ionic strength of physiological environment (Vu and Chen (2019)).

Antibodies against SARS-CoV-2 are generally detected using either the recombinant spike protein or the smaller RBD portion of the spike protein. This study only tested detected binding of antibodies to RBD but similar results would be expected with the recombinant spike protein as well (Premkumar et al. (2020)). Coating the wells with anti-SARS-CoV-2 antibodies instead of spike RBD antigen may enable rapid EIS detection of viral particles in patient samples, although further testing is needed to determine the limit of detection for that approach. There is a need for rapid patient testing due to the highly contagious nature of SARS-CoV-2 particularly when compared to viruses from the same family including, Middle East respiratory syndrome coronavirus (MERS-CoV) (fatality rate 34%) and severe acute respiratory syndrome coronavirus (SARS-CoV) (mortality rate 11%).

This study demonstrated the feasibility of using a quantitative EIS technique with commercially available equipment for rapid and accurate detection of SARS-CoV-2 antibodies at clinically relevant concentrations. This approach could enable rapid screening of patient samples, expanded serological surveys to assess anti-SARS-CoV-2 antibody levels in the community, and potentially enhance assessment of vaccine activity.

## Data Availability

The impedance data is available upon request.

## Acknowledgements

The following reagent was produced under HHSN27220-1400008C and obtained through BEI Resources, NIAID, NIH: Spike Glycoprotein Receptor Binding Domain (RBD) from SARS-Related Coronavirus 2, Wuhan-Hu-1, Recombinant from HEK293 Cells, NR-52306.

---

**Figure S1:**
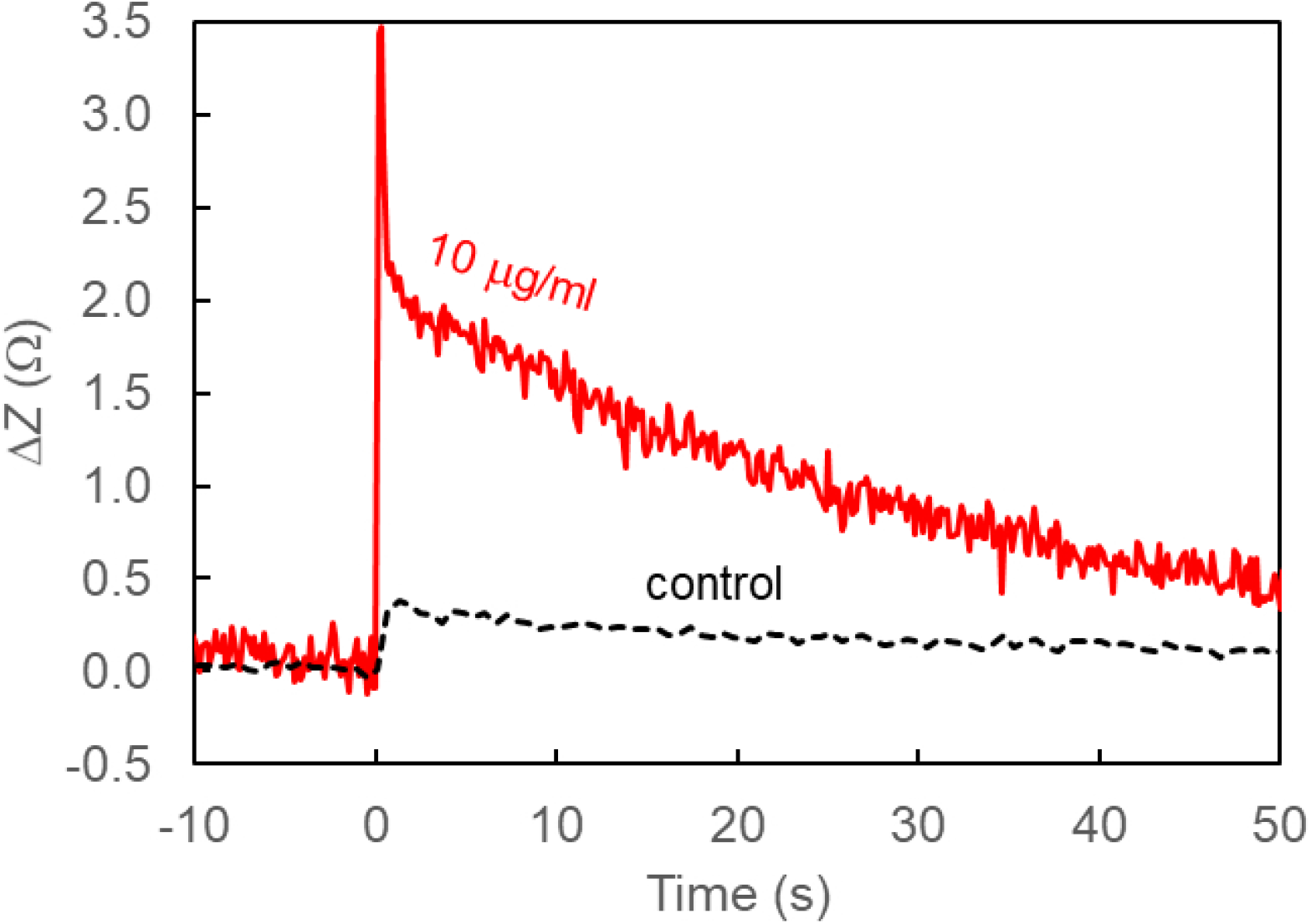
Measurements with greater time resolution were acquired with a different impedance analyzer (Agilent 4294A, 10 kHz, 0.5 V), demonstrating improved resolution.

## References

Berggren, C., Bjarnason, B., Johansson, G., 2001. Capacitive biosensors. Electroanalysis: An International Journal Devoted to Fundamental and Practical Aspects of Electroanalysis 13, 173–180.

Bordi, F., Cametti, C., Colby, R.H., 2004. Dielectric spectroscopy and conductivity of polyelectrolyte solutions. Journal of Physics: Condensed Matter 16, R1423.

Castiello, F.R., Porter, J., Modarres, P., Tabrizian, M., 2019. Interfacial capacitance immunosensing using interdigitated electrodes: the effect of insulation/immobilization chemistry. Physical Chemistry Chemical Physics 21, 15787–15797.

Cesewski, E., Johnson, B.N., 2020. Electrochemical biosensors for pathogen detection. Biosensors and Bioelectronics, 112214.

Choi, J.R., 2020. Development of point-of-care biosensors for covid-19. Frontiers in Chemistry 8, 517.

Daniels, J.S., Pourmand, N., 2007. Label-free impedance biosensors: Opportunities and challenges. Electroanalysis: An International Journal Devoted to Fundamental and Practical Aspects of Electroanalysis 19, 1239–1257.

Feldman, Y., Ermolina, I., Hayashi, Y., 2003. Time domain dielectric spectroscopy study of biological systems. IEEE transactions on dielectrics and electrical insulation 10, 728–753.

Gaurav, A., Shukla, P.,. Rapid detection of covid-19 causative virus (sars-cov-2) using fet-based biosensor.

Hanna, D.M., Gross, B.A., Lempicki, E., Oakley, B., Kandlikar, S., Stryker, G.A., 2006. Detection of vesicular stomatitis virus using a capacitive immunosensor, in: 2005 IEEE Engineering in Medicine and Biology 27th Annual Conference, IEEE. pp. 534-537.

Kulin, S., Kishore, R., Hubbard, J.B., Helmerson, K., 2002. Real-time measurement of spontaneous antigen-antibody dissociation. Biophysical journal 83, 1965–1973.

Law, J.W.F., Ab Mutalib, N.S., Chan, K.G., Lee, L.H., 2015. Rapid methods for the detection of foodborne bacterial pathogens: principles, applications, advantages and limitations. Frontiers in microbiology 5, 770.

Leva-Bueno, J., Peyman, S.A., Millner, P., 2020. A review on impedimetric immunosensors for pathogen and biomarker detection. Medical Microbiology and Immunology.

Li, Q., Guan, X., Wu, P., Wang, X., Zhou, L., Tong, Y., Ren, R., Leung, K.S., Lau, E.H., Wong, J.Y., et al., 2020. Early transmission dynamics in wuhan, china, of novel coronavirus-infected pneumonia. New England Journal of Medicine.

Liu, X., Jiang, H., 2017. Construction and potential applications of biosensors for proteins in clinical laboratory diagnosis. Sensors 17, 2805.

Luka, G., Samiei, E., Dehghani, S., Johnson, T., Najjaran, H., Hoorfar, M., 2019. Label-free capacitive biosensor for detection of cryptosporidium. Sensors 19, 258.

Premkumar, L., Segovia-Chumbez, B., Jadi, R., Martinez, D.R., Raut, R., Markmann, A., Cornaby, C., Bartelt, L., Weiss, S., Park, Y., et al., 2020. The receptor binding domain of the viral spike protein is an immunodominant and highly specific target of antibodies in sars-cov-2 patients. Science Immunology 5.

Quoc, T.V., Wu, M.S., Bui, T.T., Duc, T.C., Jen, C.P., 2017. A compact microfluidic chip with integrated impedance biosensor for protein preconcentration and detection. Biomicrofluidics 11, 054113.

Saylan, Y., Erdem, O., Unal, S., Denizli, A., 2019. An alternative medical diagnosis method: Biosensors for virus detection. Biosensors 9, 65.

Seo, G., Lee, G., Kim, M.J., Baek, S.H., Choi, M., Ku, K.B., Lee, C.S., Jun, S., Park, D., Kim, H.G., et al., 2020. Rapid detection of covid-19 causative virus (sars-cov-2) in human nasopharyngeal swab specimens using field-effect transistor-based biosensor. ACS nano 14, 5135–5142.

Sondag-Huethorst, J., Fokkink, L., 1995. Electrochemical characterization of functionalized alkanethiol monolayers on gold. Langmuir 11, 22372241.

Świetlow, A., Skoog, M., Johansson, G., 1992. Double-layer capacitance measurements of self-assembled layers on gold electrodes. Electroanalysis 4, 921–928.

Teeparuksapun, K., Hedström, M., Kanatharana, P., Thavarungkul, P., Mattiasson, B., 2012. Capacitiveimmunosensorforthedetectionofhostcell proteins. Journal of biotechnology 157, 207–213.

Vu, C.A., Chen, W.Y., 2019. Field-effect transistor biosensors for biomedical applications: recent advances and future prospects. Sensors 19, 4214.

Wang, L., Filer, J.E., Lorenz, M.M., Henry, C.S., Dandy, D.S., Geiss, B.J., 2019. An ultra-sensitive capacitive microwire sensor for pathogen-specific serum antibody responses. Biosensors and Bioelectronics 131, 46–52.

Wang, L., Veselinovic, M., Yang, L., Geiss, B.J., Dandy, D.S., Chen, T., 2017. A sensitive dna capacitive biosensor using interdigitated electrodes. Biosensors and Bioelectronics 87, 646–653.

